# Height loss in adulthood is associated with health outcomes in later life in men and women enrolled in the 1946 UK Birth Cohort (NSHD)

**DOI:** 10.1101/2023.11.04.23298098

**Authors:** Katarina L. Matthes, Kaspar Staub

## Abstract

**Objective:** To investigate the relationship between height in childhood and relative height loss in adulthood, and to examine the association between height loss and health at age 69.

**Design:** Cohort study.

**Setting:** Data from one of the oldest ongoing cohort studies, the National Survey of Health and Development (NSHD, 1946 UK birth cohort)

**Participants:** 2,119 study participants who completed the nurse home visit during the 24^th^ and most recent available follow-up examination at age 69.

**Main outcome measures:** Linear regression models to estimate the association between measured height in childhood years relative height loss between ages 36 and 69. Logistic regression models using generalized additive models to estimate the probability of worse health at age 69 (chronic disease score, general health status, osteoarthritis, and pain while walking) in association with height loss.

**Results:** Between the ages of 36 and 69, men lost an average of 2.03 cm and women 2.44 cm. Women lost significantly more height than men (p<0.001). The taller the participants were at a young age, the more height they lost in adulthood. There was a significant association between height loss in adulthood on the one hand and general health, chronic disease score (in men), osteoarthritis (in men), and walking pain at age 69. These findings largely persisted after adjusting the models for overweight, sociodemographic information, and lifestyle factors earlier in life.

**Conclusions:** Height loss seems to mirror deteriorating health as people age. Height measurement and assessment of height loss should be part of regular examinations after the age of 40 to monitor general health status, especially in the case of severe height loss.

## 1 Introduction

Human body height is not a constant, but is subject to dynamic processes of change throughout a person’s lifetime [1,2]. Height increases as a product of physical growth until adulthood, then is more or less stable between the ages of 20 and about 40, after which most people lose height as they biologically age [3]. Height loss typically begins between the ages of 40 and 45, and women are usually more affected than men [4]. However, the extent of height loss (between 1.5 cm and 5 cm between the ages of 40 and 80) varies between studies and is probably related to differences in sample composition. Loss of height may be caused by various health conditions such as spinal changes (e.g., vertebral fractures, muscle weakness, postural changes, disc degeneration, spinal deformities, kyphosis, degenerative lumbar scoliosis) [5].

Several associations of height loss with health outcomes have been found using longitudinal data: The Framingham Study found an association between height loss and hip fracture in men [5], as did the EPIC-Norfolk population cohort study [6]. In postmenopausal women, associations were found between height loss and fracture risk and functional status [7]. The Hiroshima Cohort Study documented associations between height loss and health-related quality of life [8]. It has also been shown that the risk of all-cause mortality was higher in men with a height loss of >=3 cm [9]. In addition, the Pyeongchang Rural Area Cohort in South Korea has shown that greater height loss is associated with a higher risk of frailty and sarcopenia [10].

However, longitudinal studies over 30 years or more of potential height loss in adulthood that allow comprehensive assessment of later life outcomes are rare. And even fewer attempts have been made to look at long-term trajectories of height and to relate growth to peak adult height to adult height loss and health in later life, largely because the data base for this question has been lacking. Overall, it can be said that there is not yet enough evidence on the association between either growth or height loss (or both in combination) and health in later life. Both growth and height loss are potential risk factors for health in later life that are common to most people. Better assessment of the evidence is therefore an important public health desideratum.

This paper addresses these research gaps using a unique data source from one of the oldest ongoing cohort studies: The National Survey of Health and Development (NSHD) or 1946 British Birth Cohort, whose participants can now be followed in detail from birth to almost 70 years of age. The aim of this study was to examine 1) the extend of height loss in the 1946 UK birth cohort and its association with height in childhood, 2) the association between height loss in adulthood and health status at age 69.

## 2 Materials and methods

### The NSHD study

The National Survey of Health and Development (NSHD) or 1946 British Birth Cohort is the longest running and oldest of the British birth cohort studies [11]. The NSHD cohort (initially n=5,362) is a nationally representative sample of the larger cross-sectional Maternity Survey, which was designed to investigate the reasons for the low birth rate in the United Kingdom by studying all n=16,695 births that occurred in England, Wales, and Scotland during one week in March 1946. Following the success of the Maternity Survey, the NSHD was designed as a follow-up study to examine the health and development of about one-third of the babies. As of 2015, cohort participants have been followed up 24 times. The range of data generated is very extensive, with a total of about 25,000 variables available for research. Height was measured at least 10 times after birth, at ages 2, 4, 6, 7, 11, 15, 36, 43, 53, 63, and 69. Growth patterns, especially during puberty, have been described in detail for the NSHD [12].

The 24th follow-up in 2015 was the most recent follow-up and included height measurement, a postal questionnaire at age 68, and a home visit at age 69 that focused on morbidity and its consequences, as well as repeated functional and mental health assessments and updated reports on health and living conditions [13]. The target sample consisted of 2,816 study participants still living in the UK. Of the remaining 2,546 (47%) participants, 957 (18%) had died, 620 (12%) had previously withdrawn permanently, 574 (11%) were living abroad, and 395 (7%) had been lost to follow-up for more than 5 years. Overall participation and home visit participation remained high (94 and 80%, respectively) [13].

The home visit by a research nurse took place at age 69 for the majority (97%). A total of 2,149 study participants completed the home visit. During the home visit (described in detail on the NSHD data portal [14]), nurses administered an electronic questionnaire asking about health, disability, medications, smoking, and exercise habits. The nurses took measurements of height, blood pressure, grip strength, and respiration, and also administered tests of balance and walking speed. During the home visit, the study participant was also asked to complete a paper questionnaire, the short (28-item) General Health Questionnaire.

### Selected variables

#### Health outcome variables: Health at age 69

The following variables were selected as outcome variables A) The chronic disease score, which is based on the total 19-point physician-diagnosed chronic disease score. Two groups were classified: “<3 points” vs. “>=3 points”. B) Based on a score for multiple symptoms, participants were assigned to four summary health groups: “no illness, no symptoms”, “healthier middle”, “less healthy middle”, and “multiple illnesses and symptoms”. For this study, the first and second groups were combined, and the third and fourth groups were combined. C) Physical ability and mobility were included by the question “Does pain limit your ability to walk? (“yes” vs. “no”). D) Because of the well-documented association between height loss and bone health, the question “Have you been diagnosed with osteoarthritis? (“yes” vs. “no”) was included.

#### Primary exposure variable: Height

Height measurements at age 69, during the 2015 nurse home visit, followed a detailed manual for nurses (published elsewhere [15]). The same procedure was used for height measurements at younger ages. At ages 20 and 26, height was self-reported. Therefore, information at ages 20 and 26 was not considered in this study. For height measurements at ages 2, 4, 7, 11, and 15, measurements were taken as imperials and recorded to the nearest whole inch. Height measurements at age 36 and later were taken in metric to an accuracy of 1 mm (e.g., 1.814 m). To assess average heights in this study, all height measurements in inches up to age 15 were also converted to centimetres (e.g., 181.4 cm). In addition, height gain between 2 and 7 years of age, between 7 and 11 years of age, and between 11 and 15 years of age was calculated. The SITAR (Superimposition by Translation And Rotation) parameter size height(pubertal height according to SITAR growth modelling [12,16]) was used to include pubertal growth in the analysis. Absolute height loss between 36 and 69 years of age was calculated as the delta in cm between the two height measurements. We excluded n=7 individuals whose height loss between 36 and 69 years was >15 cm or whose height gain was >5 cm. A height gain between 0 cm and 5 cm was considered as a height loss of 0 cm for the analysis (Supplement Figure 1). Relative height loss was expressed as the percentage of height loss compared to height at age 36.

#### Sociodemographic and lifestyle aspects

The following variables, which are known to be associated with either height or health in old age, were selected as control variables: Body mass index (BMI) at age 36 years was calculated from measured height and weight (BMI=weight in kg divided by the square of height in m) and then categorized as a binary variable “overweight” (BMI>=25.0kg/m2) vs. “normal weight” (BMI<25.0kg/m2). As an indicator of socioeconomic position (SEP) in adulthood, the highest educational attainment up to the age of 43 years was used. The subgroups “A-level or equivalent” and “higher” were binary coded as “higher education”, all other subgroups (“O-level or equivalent”, “vocational only”, “no qualifications”) were coded as “no higher education”. As a proxy for childhood SEP, “childhood social class” was used as a summary variable. The subgroups “partly skilled” and “unskilled” were combined into “low SEP”, “skilled (non-manual)” and “skilled (manual)” into “medium SEP”, and “professional etc.” and “medium” into “high SEP”. The standard regions were used as a rough regional variable for the region of residence at the time of birth and classified into three categories: North (Scotland, Northern, Yorkshire/Humberside, Northwest England), Middle (Wales, East Midlands, West Midlands, East Anglia) and South (South East England, South West England). The selection of lifestyle aspects with sufficient coverage in the data set was rather modest. Rough indicators for the period up to age 69 were included: First, from the 2009 survey at age 63, the question “Have you ever smoked cigarettes regularly?”, coded “yes” or “no”. Second, as a proxy for healthy eating and also from the 2009 survey round, the question “How often do you eat fruit?” was coded into “not every day” and “every day”. And as an indicator of physical activity, the summary variable “Exercise at age 29-49”, whose responses were again coded into two groups: “<1 per week” and “>=1 per week”.

### Statistics

All analyses were performed separately for each sex. Linear regression was used to examine the association of height in earlier years with subsequent relative height loss. For this purpose, height at ages 2, 7, 11, 15, and 36, height differences between 2 and 7, 7 and 11, and 11 and 15, and SITAR size height were used as explanatory variables and analysed in separate models. Each of these variables was z-transformed to ensure comparability between models. Univariate analysis was performed first, followed by multivariate analysis including the aforementioned sociodemographic and lifestyle factors.

The probability of the four binary response variables (chronic disease score, summary health groupings, osteoarthritis, pain while walking) and relative height loss were estimated by logistic regression models using generalized additive models (GAM). GAMs can fit nonlinear relationships between response and exploratory variables (in addition to linear relationships). First, the probability of each outcome was estimated univariately, considering only relative height loss as a nonlinear term. Second, three models were estimated for each outcome, stepwise adding different explanatory variables, and using the sample size of the fully adjusted third model. In the first model, the probability of each outcome was again estimated univariately. In the second model, “SITAR height size” was included as a non-linear term (to control for pubertal height) and socio-demographic factors were included as linear terms. In the third model, lifestyle factors were additionally included as linear terms. All statistical analyses were performed with R version 4.2.2. The R package “tidyverse” [17] was used for data manipulation and to generate all figures. The “mgcv” package [18] was used for the generalized additive models (GAM). All code is available in the repository at https://github.com/KaMatthes/height_loss

### Data availability and ethics

The data owner (MRC Unit for Lifelong Health and Ageing at UCL (LHA)) actively encourages the sharing of NSHD data with researchers. A data sharing and confidentiality agreement has been signed between the LHA and the University of Zurich, and the present project was approved by the NSHD Data Sharing Committee after submission of a short protocol. A subset of the 1946 to 2018 data was delivered to the study team in August 2021 [19–21]. Detailed information on participation and ethics can be found on the NSHD study website [22]. All study participants are volunteers and have/had the right to withdraw from the study at any time without giving a reason. For the 2015 data collection at age 69, the NSHD study team received ethical approval from the NRES Queen Square REC (14/LO/1073) and the Scotland A REC (14/SS/1009) [13]. This project has also been approved by the LSHTM MSc Ethics Committee (ref: 25746, approved on August 3, 2021).

## 3 Results

Table 1 shows descriptive results for the full sample and the 2015 nurse sample. A total of 2119 participants (1038 men and 1081 women) completed the nurse visit at home during the examination at age 69 in 2015, and information on height at ages 36 and 69 was available. At age 36, men were on average 175.6 cm (SD 6.4) tall, and women 162.9 cm (SD 6.4) tall. At age 69, 33 years later, men were 173.9 cm (SD 6.5) and women 160.6 cm (SD 5.9) tall. This results in a mean height loss of 2.03 cm (SD 1.79) for men (1.15% (SD 1.00%) of the height at age 36) and a height loss of 2.44 cm (SD 2.03) for women (1.50% (SD 1.23%) of the height at age 36). Women lost significantly more height than men (two-tailed, two-sample t-test p < 0.001). Comparing the mean height at ages 2 to 69 years of the 2015 nurse visit participants with the mean values for the respective total cohort (Supplementary Figure 2), the differences between the mean heights are very small. The density of heights at ages 26, 43, 53, and 69 years (Supplementary Figure 3) shows that the loss of height affected the entire height distributions, which were shifted to the left on the x-axis, starting with the distributions at age 53 years.

**Table 1:** Descriptive statistics for the 2015 nurse visit sample at age 69 (left) and the full cohort since 1946 (right).

|  |  | Nurse visit at age 69 |  |  |  | Full Sample |  |  |  |
| --- | --- | --- | --- | --- | --- | --- | --- | --- | --- |
|  |  | Male |  | Female |  | Male |  | Female |  |
|  |  | n | mean(sd) | n | mean (sd) | n | mean (sd) | n | mean (sd) |
| Height in cm at age 2 | Mean (SD) | 856 | 86 (5.1) | 861 | 84.9 (4.5) | 2135 | 85.8 (5.2) | 1871 | 84.7 (4.8) |
| Height in cm at age 7 | Mean (SD) | 901 | 120.5 (5.4) | 940 | 120.1 (5.5) | 2145 | 120.4 (5.6) | 1988 | 119.5 (5.7) |
| Height in cm at age 11 | Mean (SD) | 882 | 141.1 (6.6) | 908 | 141.6 (6.9) | 2080 | 140.7 (6.7) | 1908 | 140.9 (7.1) |
| Height in cm at age 15 | Mean (SD) | 818 | 162.4 (8.8) | 841 | 159.1 (6.1) | 1911 | 162.0 (8.9) | 1721 | 158.4 (6.3) |
| Height in cm at age 36 | Mean (SD) | 932 | 175.6 (6.4) | 990 | 162.9 (6.4) | 1635 | 175.3 (6.6) | 1649 | 162.3 (6.1) |
| Height in cm at age 69 | Mean (SD) | 1038 | 173.9 (6.5) | 1081 | 160.6 (5.9) | 1038 | 173.9 (6.5) | 1081 | 160.6 (5.9) |
| Height loss in cm<br>(age 36-69) | Mean (SD) | 1038 | 2.03 (1.79) | 1081 | 2.44 (2.03) | 1038 | 2.03 (1.79) | 1081 | 2.44 (2.03) |
| Relative height loss in %<br>(age 36-69) | Mean (SD) | 1038 | 1.15 (1.00) | 1081 | 1.50 (1.23) | 1038 | 1.15 (1.00) | 1081 | 1.50 (1.23) |
|  |  | n | % | n | % | n | % | n | % |
| Education as adults<br>(at age 43) | No higher education | 505 | 49.0 | 672 | 62.7 | 1403 | 58.4 | 1569 | 70.8 |
|  | Higher education | 526 | 51.0 | 399 | 37.3 | 999 | 41.6 | 648 | 29.2 |
|  | Total | 1031 | 100.0 | 1071 | 100.0 | 2402 | 100.0 | 2217 | 100.0 |
| Region of birth<br>(at age 0) | South | 423 | 40.8 | 453 | 41.9 | 1026 | 36.5 | 989 | 38.9 |
|  | Middle | 248 | 23.9 | 273 | 25.3 | 666 | 23.7 | 626 | 24.6 |
|  | North | 367 | 35.4 | 355 | 32.8 | 1121 | 39.9 | 927 | 36.5 |
|  | Total | 1038 | 100.0 | 1081 | 100.0 | 2813 | 100.0 | 2542 | 100.0 |
| Overweight<br>(at age 36) | No | 548 | 58.9 | 757 | 77.9 | 912 | 56.0 | 1200 | 74.4 |
|  | Yes | 382 | 41.1 | 215 | 22.1 | 717 | 44.0 | 413 | 25.6 |
|  | Total | 930 | 100.0 | 972 | 100.0 | 1629 | 100.0 | 1613 | 100.0 |
| SEP in childhood<br>(at age 0-2) | Low SEP | 295 | 29.8 | 304 | 30.0 | 620 | 25.3 | 570 | 25.7 |
|  | Medium SEP | 482 | 48.7 | 483 | 47.6 | 1192 | 48.7 | 1089 | 49.1 |
|  | High SEP | 213 | 21.5 | 228 | 22.5 | 638 | 26.0 | 557 | 25.1 |
|  | Total | 990 | 100.0 | 1015 | 100.0 | 2450 | 100.0 | 2216 | 100.0 |
| Fruit consumption<br>(at age 63) | Not every day | 289 | 33.3 | 236 | 24.5 | 367 | 34.9 | 289 | 25.0 |
|  | Ever day | 580 | 66.7 | 729 | 75.5 | 684 | 65.1 | 866 | 75.0 |
|  | Total | 869 | 100.0 | 965 | 100.0 | 1051 | 100.0 | 1155 | 100.0 |
| Ever smoked<br>(at age 63) | No | 361 | 45.9 | 520 | 61.4 | 467 | 46.6 | 673 | 62.0 |
|  | Yes | 426 | 54.1 | 327 | 38.6 | 535 | 53.4 | 413 | 38.0 |
|  | Total | 787 | 100.0 | 847 | 100.0 | 1002 | 100.0 | 1086 | 100.0 |
| Physical activity<br>(at age 29-49) | <1 / week | 409 | 49.7 | 591 | 67.3 | 409 | 49.6 | 596 | 67.5 |
|  | >=1 / week | 414 | 50.3 | 287 | 32.7 | 416 | 50.4 | 287 | 32.5 |
|  | Total | 823 | 100.0 | 878 | 100.0 | 825 | 100.0 | 883 | 100.0 |
| General health status<br>(at age 69) | Healthy | 722 | 77.5 | 640 | 64.4 |  |  |  |  |
|  | Less/not healthy | 210 | 22.5 | 354 | 35.6 |  |  |  |  |
|  | Total | 932 | 100.0 | 994 | 100.0 |  |  |  |  |
| Chronic disease score<br>(at age 69) | <3 | 833 | 80.5 | 851 | 78.9 |  |  |  |  |
|  | >=3 | 202 | 19.5 | 227 | 21.1 |  |  |  |  |
|  | Total | 1035 | 100.0 | 1078 | 100.0 |  |  |  |  |
| Osteoarthritis<br>(at age 60-69) | No | 869 | 83.7 | 803 | 74.5 |  |  |  |  |
|  | Yes | 169 | 16.3 | 275 | 25.5 |  |  |  |  |
|  | Total | 1038 | 100.0 | 1078 | 100.0 |  |  |  |  |
| Pain while walking<br>(at age 69) | No | 714 | 80.3 | 695 | 72.5 |  |  |  |  |
|  | Yes | 175 | 19.7 | 263 | 27.5 |  |  |  |  |
|  | Total | 889 | 100.0 | 958 | 100.0 |  |  |  |  |

The distribution of region of birth and SEP in childhood was similar for men and women. However, women were less likely to have higher education as adults, while men were more likely to be overweight and to smoke at age 36. In contrast, between the ages of 29 and 49, men were more likely to exercise, while women were more likely to eat fruit. When comparing the distribution of sociodemographic and lifestyle variables for participants who completed the nurse visit at age 69 with the entire cohort, only minor differences emerged. Participants in the 2015 nurse visit were slightly better educated, more likely to be born in the South, less likely to be overweight at age 36, and more likely to have a higher childhood SEP than the entire cohort. In terms of health outcomes at age 69, women generally scored slightly worse than men: Women were more likely to have a “less/not healthy” summary health status (35.6% vs. 22.5% for men, p<0.001), slightly more likely to have osteoarthritis (25.5% vs. 16.3% for men, p<0.001), and more likely to have pain when walking (27.5% vs. 19.7% for men, p<0.001). The proportions were similar for the chronic disease score (21.1% in women vs. 19.4% in men, p=0.410).

Figure 1 shows the coefficients in z-values and 95% confidence intervals (CI) of each linear regression model. It is evident that the taller men and women are at each age, the greater the relative loss in height by age 69, although this is not significant at all ages. After adjustment, height at ages 7 (0.11 (95% CI 0.02-0.21)), 15 (0.10 (95% CI 0.00-0.20)), and 36 (0.18 (95% CI 0.09-0.27)) still plays an important role in height loss later in life for men (Supplementary Table 1), as does SITAR height at puberty (0.13 (95% CI 0.04-0.22)). In women, only height at 36 years is significantly associated with relative height loss (0.18 (95% 0.07-0.28)). Greater height gain between ages is not significantly associated with greater relative height loss.

**Figure 1:**
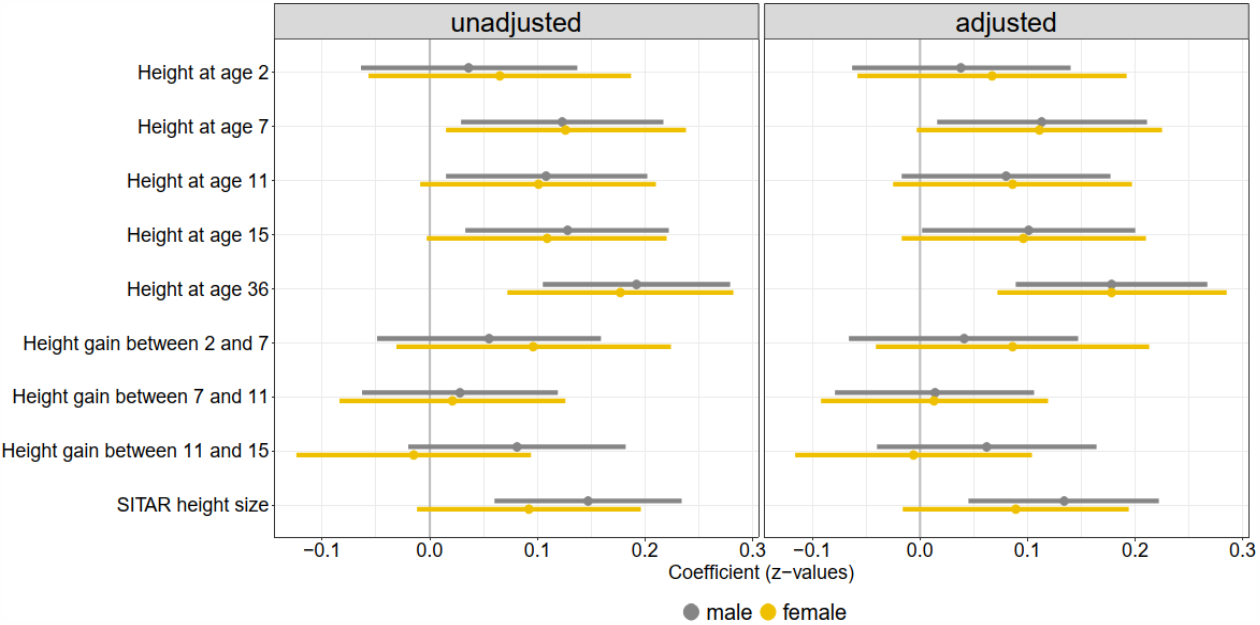
Coefficients (z-values) of linear regression models estimating the effect of relative height loss on various height parameters in childhood and at age 36 years. Unadjusted and adjusted for overweight, education, region of birth, SEP in childhood, smoking, fruit consumption and physical activity.

For both sexes, the probability of a worse health outcome increases significantly with increasing relative height loss (Figure 2, detailed regression results in Supplementary Tables 2-5). The highest probability is seen for general health, followed by pain when walking. For all four health outcomes, women are more likely than men to be in worse health with increasing height loss. When the model is adjusted for socio-demographic (model 2) and lifestyle factors (model 3) and compared with the univariate model 1, the association between height loss and health outcomes persists but is slightly mitigated (Figures 3 and 4, detailed regression results in Supplementary Tables 2-5). In addition to height loss, being over-weight at age 36 is the most important factor in poorer health outcomes at age 69 and is associated with poorer general health (model 3 men: OR 1.97 (95% CI 1.25-3.1), women: OR 1.62 (95% CI 1.04-2.52)), with the presence of osteoarthritis (model 3 men: OR 2.59 (95% CI 1.51-4.45), women: OR 1.61 (95% CI 1.02-2.55)), with a higher chronic disease score in men (model 3, OR 2.89 (95% CI 1.75-4.76)) and a higher risk of walking pain in women (model 3, OR 2.86 (95% CI 1.78-4.58)).

**Figure 2:**
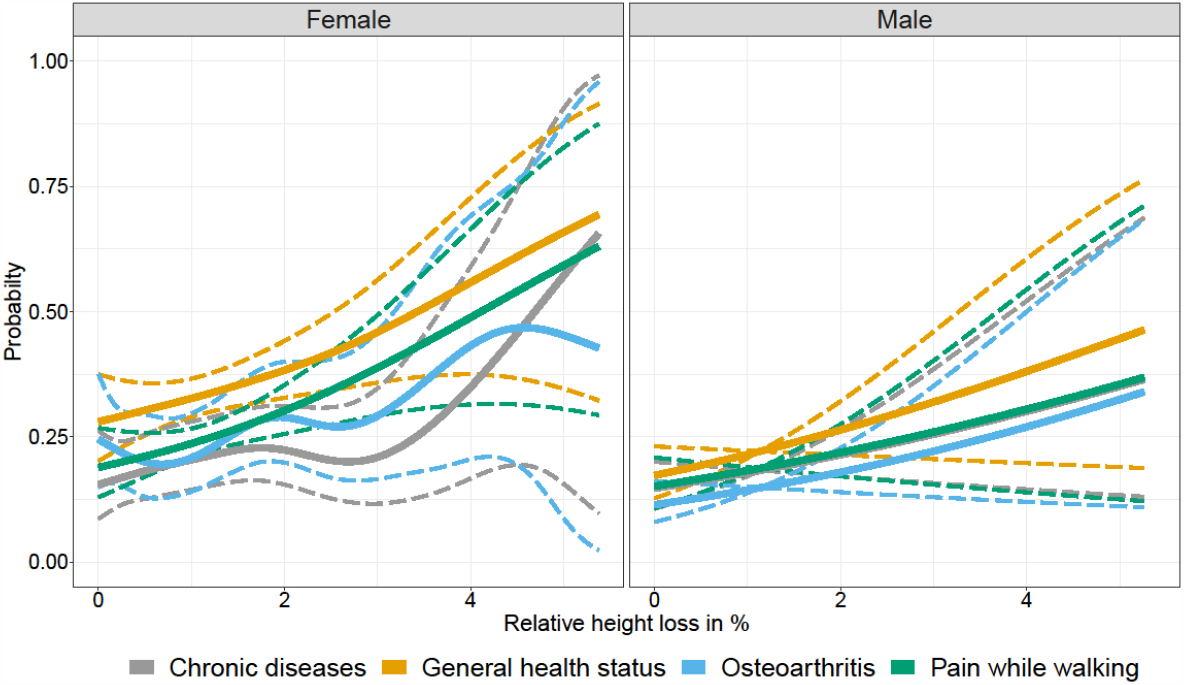
Probabilities from univariate logistic GAMs of the four health outcome variables associated with height loss in men and women using the full sample size.

**Figure 3:**
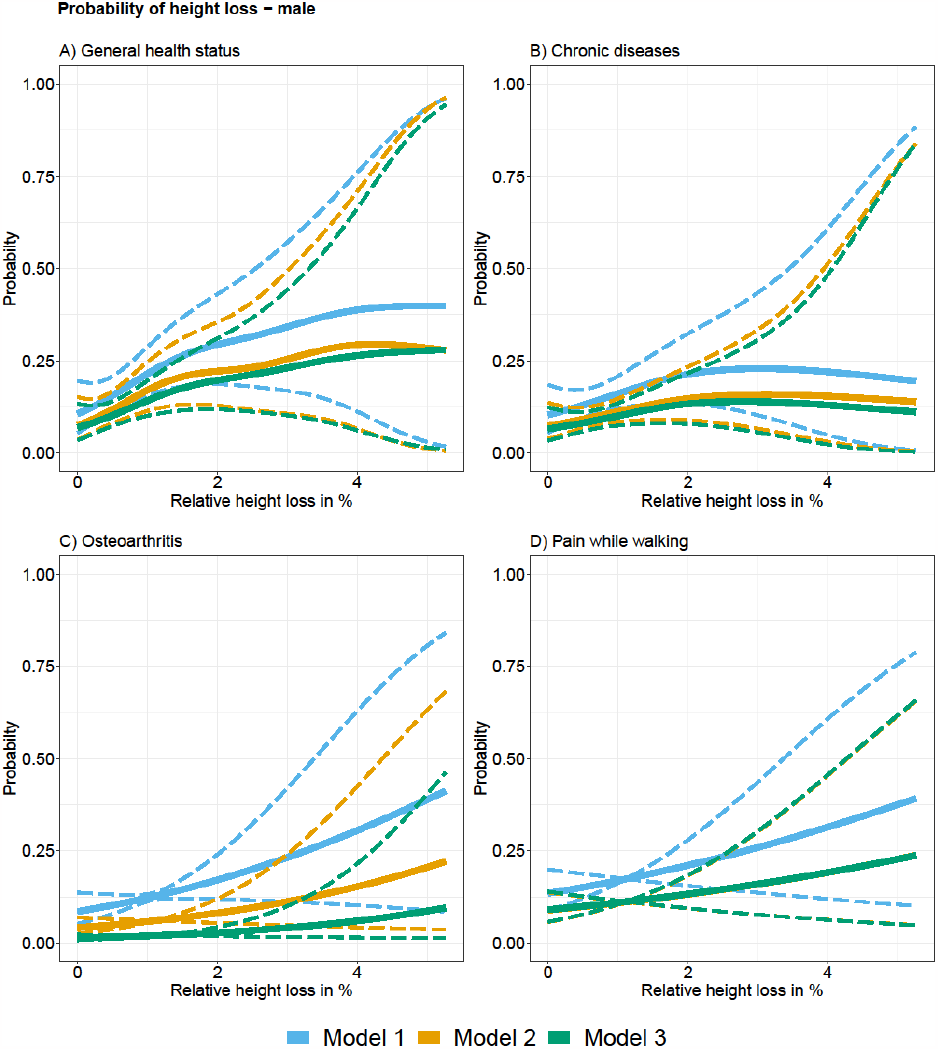
Probabilities of logistic GAMs of health in later life and height loss in men using the reduced sample size (complete cases). Model 1: univariate: only relative height loss, Model 2: multivariate: adjusted for SITAR height for overweight, education, region of birth, SEP in childhood, Model 3: additionally adjusted for smoking, fruit consumption and physical activity.

**Figure 4:**
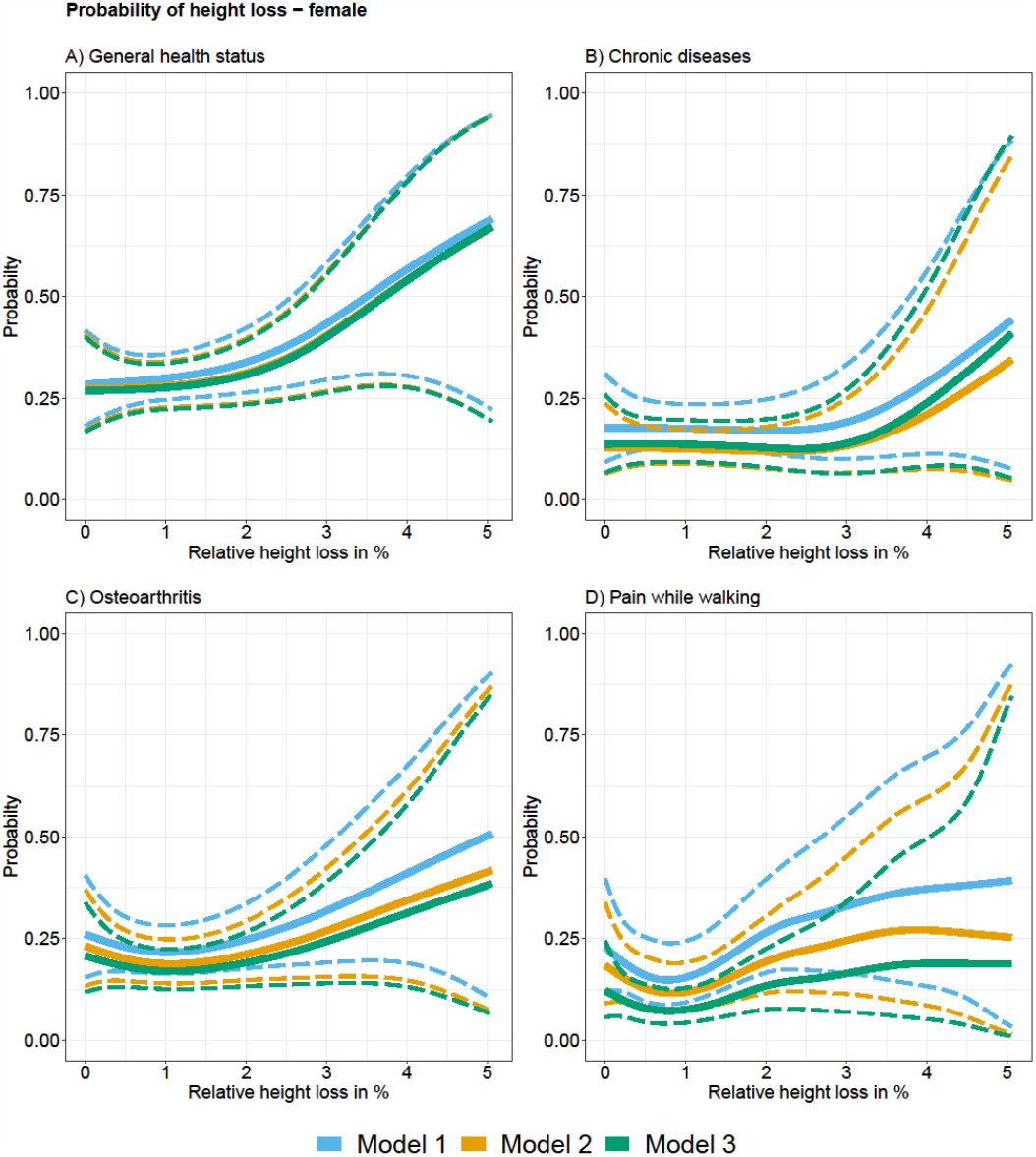
Probabilities of logistic GAMs of health in later life and height loss in women using the reduced sample size (complete cases). Model 1: univariate: only relative height loss, Model 2: multivariate: adjusted for SI-TAR height for overweight, education, region of birth, SEP in childhood, Model 3: additionally adjusted for smoking, fruit consumption and physical activity.

## 4 Discussion

This study examined the oldest birth cohort study in the United Kingdom, the NSHD or 1946 UK Birth Cohort. The focus was on the association between lifetime changes in height and adult height loss on the one hand, and health status at age 69 on the other. It was shown that between the ages of 36 and 69 (over 33 years), men lost an average of 2.03 cm (1.15%) of their peak adult height and women lost 2.44 cm (1.50%). The magnitude of height loss in the NSHD over the 33 years between ages 36 and 69 is rather small compared to the limited literature available. In other long-term studies, height loss ranged from 1.5 cm to 4 cm per decade of follow-up [5,23]. However, it must be emphasized that none of these other studies represented a similarly homogeneous cohort. In most cases, participants were recruited on a rolling basis over a longer period of time and in late adulthood, and follow-up was shorter. In addition, none of these other studies was designed to be a representative cohort since birth. It should also be emphasized that the NSHD participants are now “only” about 70 years old. It is known from other studies that height loss increases with age and is particularly pronounced between the ages of 70 and 80 [4].

The results presented here are consistent with previous literature that excessive height loss in particular increases later health risk [8,9], and that women are generally more affected by height loss than men [4,5,7,23]. This latter phenomenon may be influenced by menopause (possibly hormonal influences?) or by the generally higher risk of poorer bone health in women later in life [23]. Reasons for height loss include muscle weakness, postural changes, disc degeneration, frailty, joint space narrowing, spinal deformity, and kyphosis. The extent to which women are more affected by these problems remains to be determined.

The fact that being overweight in young adulthood is an additional risk factor for poorer health outcomes in later life is supported by a previous study, also using NSHD data, which showed that the higher the BMI, the greater the risk of back pain in adulthood [24]. Weight has also been shown to be a risk factor for height loss [25]. The results presented here also confirm a recent study of the NSHD data, which found little significant association for various growth parameters in childhood with osteoarthritis at age 53 [26]. Again, childhood height parameters played only a minor role.

The strength of the present study lies in the underlying unique data source, a representative cohort, all born in March 1946 and followed up with a detailed examination battery until age 69. However, the study has several limitations: First, there is selection bias in the follow-up examinations, especially later in life. It is possible that the study participants who participated in the follow-up examinations at age 69 were healthier than those who died or refused to participate (healthy participant bias). This may be an additional reason why height loss was slightly lower in the NSHD than in other studies. It has also been shown that loss to follow-up is a particular challenge in long-term cohorts, as permanent withdrawals and non-participation may increase with advancing age [13]. Second, there may have been measurement and reporting errors in height. This cannot be excluded, especially for the few participants whose height was significantly higher at age 69 than at age 36. The measurements were taken by specially trained study nurses according to a very detailed protocol (which was identical for all previous measurements), which should reduce such inaccuracies to an acceptable minimum. Now that this study has shown that height loss may be an important issue, special care should be taken to ensure that the measurement protocol is strictly followed at the next follow-up visits. An additional inaccuracy is that height is generally slightly higher in the morning than in the evening (up to a few centimetres). There is no information in the dataset about the time of day when a measurement was taken. Therefore, it is not possible to correct for this. Third, the lifestyle factors available in the NSHD data for surveys and interviews prior to 2015 were rather vague (dietary patterns) and/or often incomplete. More precise information on diet, physical activity, and smoking/alcohol over the entire life course up to the age of 69 years would allow more conclusive statements on the association with health status and confounding. Fourth, four outcome variables were selected for this study from a large catalogue of information on many diseases and symptoms at age 69. Future studies should test hypotheses more broadly in this regard.

Fifth, and almost most important, this study has shown associations between height loss and selected aspects of health in older age. Causality cannot be established. In particular, it is possible that the direction of the effect runs both ways: on the one hand, height loss could be one of several causes of poorer health in older age, but on the other hand, height loss could also be a by-product of poorer health in older age, mainly due to other causes. This means that height loss could also be a marker of underlying poor health for other reasons, i.e., a confounder rather than a predictor. To separate this sufficiently well would require even larger sample sizes, consideration of all available information on health status at all previous examinations of the participants, and possibly other study designs (e.g., Mendelian randomization, if possible).

Whether more of a cause or a consequence, height loss - especially excessive height loss - reflects deteriorating health as people age. It remains one of the great advantages of the anthropometric measure of height that it can be measured relatively easily, cheaply and reliably, even in any general practitioner’s office. If further studies confirm the relevance of height loss as a factor in later health, then height measurement and assessment of height loss should be part of regular examinations from the age of 40 to 50 to monitor general health status, especially in the case of severe height loss, and more so in women than in men. It will be intriguing in the future to follow this unique NSHD cohort and its participants as they live into their 80s.

## Acknowledgements

The authors thank Mark Tylor for support and comments, the MRC Unit for Lifelong Health and Ageing at UCL (LHA) for support and access to data, and the study team and especially the participants in the NSHD study.

## References

1 Eveleth PB, Tanner JM. Worldwide variation in human growth. 2nd ed. Cambridge <etc.>: Cambridge University Press 1990.

2 Tanner JM. Growth at Adolescence. 2nd ed. Oxford: Blackwell1962.

3 Bogin B. Patterns of Human Growth. Cambridge University Press 2008.

4 Sorkin JD, Muller DC, Andres R. Longitudinal Change in Height of Men and Women: Implications for Interpretation of the Body Mass Index: The Baltimore Longitudinal Study of Aging. Am J Epidemiol. 1999;150:969–77.

5 Hannan MT, Broe KE, Cupples LA, et al. Height loss predicts subsequent hip fracture in men and women of the Framingham Study. J Bone Miner Res. 2012;27:146–52.

6 Moayyeri A, Kaptoge S, Luben RN, et al. Estimation of absolute fracture risk among middleaged and older men and women: the EPIC-Norfolk population cohort study. Eur J Epidemiol. 2009;24:259–66.

7 Pluskiewicz W, Adamczyk P, Drozdzowska B. The significance of height loss in postmenopausal women. The results from GO Study. Int J Clin Pract. 2021;75. doi: 10.1111/ijcp.14009

8 Masunari N, Fujiwara S, Nakata Y, et al. Historical height loss, vertebral deformity, and health-related quality of life in Hiroshima cohort study. Osteoporos Int. 2007;18:1493–9.

9 Wannamethee SG. Height Loss in Older Men. Arch Intern Med. 2006;166:2546.

10 Ji S, Lee E, Kim BJ, et al. Height loss as an indicator of ageing through its association with frailty and sarcopenia: An observational cohort study. Arch Gerontol Geriatr. 2023;110:104916.

11 Wadsworth M, Kuh D, Richards M, et al. Cohort Profile: The 1946 National Birth Cohort (MRC National Survey of Health and Development). Int J Epidemiol. 2006;35:49–54.

12 Kuh D, Muthuri SG, Moore A, et al. Pubertal timing and bone phenotype in early old age: findings from a British birth cohort study. Int J Epidemiol. 2016;dyw131.

13 Kuh D, Wong A, Shah I, et al. The MRC National Survey of Health and Development reaches age 70: maintaining participation at older ages in a birth cohort study. Eur J Epidemiol. 2016;31:1135–47.

14 NSHD. Skylark Home Page. https://skylark.ucl.ac.uk/Skylark/ (accessed 1 November 2023)

15 NSHD. HOME VISIT 2015 NURSE’S MANUAL OF PROCEDURE. 2015. https://skylark.ucl.ac.uk/NSHD/lib/exe/fetch.php?media=questionnaires:2015_nurse_manual.p df (accessed 1 November 2023)

16 Cole TJ, Donaldson MDC, Ben-Shlomo Y. SITAR—a useful instrument for growth curve analysis. Int J Epidemiol. 2010;39:1558–66.

17 Wickham H, Averick M, Bryan J, et al. Welcome to the Tidyverse. J Open Source Softw. 2019;4:1686.

18 Wood SN. Generalized additive models: An introduction with R, second edition. Gen Addit Model An Introd with R, Second Ed. 2017;1–476.

19 Douglas J, Wadsworth M, Kuh D. MRC NSHD 1946-2005 Data. MRC Unit for Lifelong Health and Ageing at UCL. 2015. 10.5522/NSHD/Q101

20 Kuh D, Hardy R, Richards M, et al. MRC NSHD 2006-2012 Data. MRC Unit for Lifelong Health and Ageing at UCL. 2015. 10.5522/NSHD/Q102

21 Kuh D, Hardy R, Richards M, et al. MRC NSHD 2013-2018 Data. MRC Unit for Lifelong Health and Ageing at UCL. 2018. 10.5522/NSHD/Q103

22 NSHD. NSHD. https://nshd.mrc.ac.uk/ (accessed 1 November 2023)

23 Shimizu M, Kobayashi T, Chiba H, et al. Adult spinal deformity and its relationship with height loss: a 34-year longitudinal cohort study. BMC Musculoskelet Disord. 2020;21:422.

24 Muthuri S, Cooper R, Kuh D, et al. Do the associations of body mass index and waist circumference with back pain change as people age? 32 years of follow-up in a British birth cohort. BMJ Open. 2020;10:e039197.

25 Mai X, Marshall B, Hovey KM, et al. Risk factors for 5-year prospective height loss among postmenopausal women. Menopause. 2018;25:883–9.

26 Staines KA, Hardy R, Samvelyan HJ, et al. Life course longitudinal growth and risk of knee osteoarthritis at age 53 years: evidence from the 1946 British birth cohort study. Osteoarthr Cartil. 2021;29:335–40.

