## Supplementary Material for "Height loss in adulthood is associated with health outcomes in later life in men and women enrolled in the 1946 UK Birth Cohort (NSHD)"

---

***Supplementary Figure 1:*** *Distribution of height loss in adulthood in cm.*

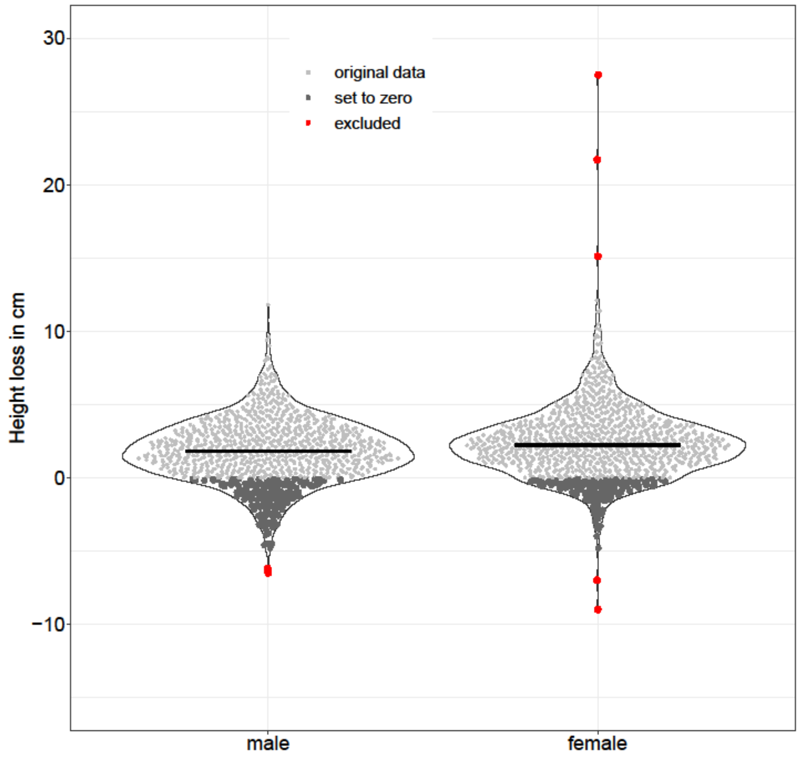

***Supplementary Figure 2:*** *Box plots of height distribution for each age of measurement.*

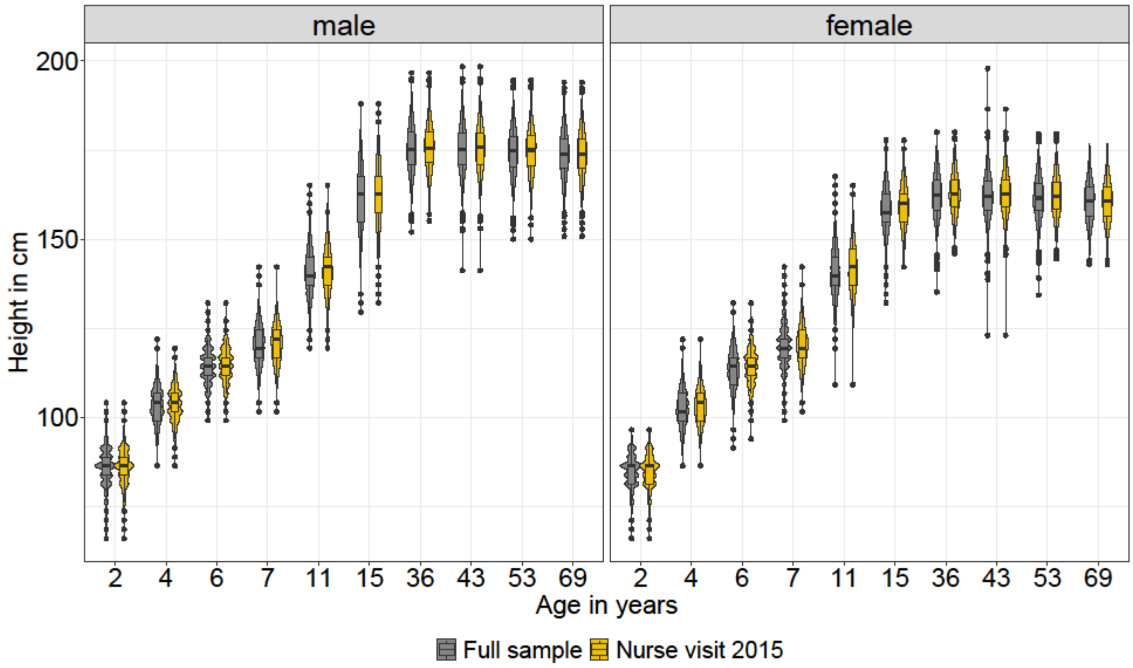

***Supplementary Figure 3:*** *Density plots of the body height distribution at all measurement points in adulthood.*

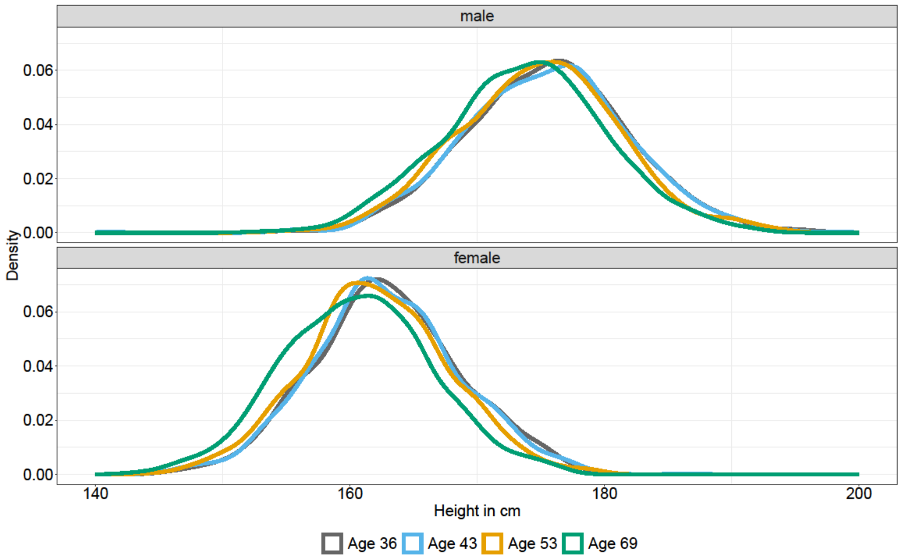

***Supplementary Table 1:*** *Coefficients (z-values) of the linear regression models to estimate the effect of relative height loss on different height parameters during childhood and age 36. Unadjusted and adjusted for overweight, education, region of birth, SEP in childhood, smoking, fruit consumption and physical activity. Grey shaded cells show significant results.*

| | Male<br>Coefficient (95% CI) | | Female<br>Coefficient (95% CI) | |
|---|---|---|---|---|
| | **unadjusted** | **adjusted** | **unadjusted** | **adjusted** |
| **Height at age 2** | 0.04 (-0.06 to 0.14) | 0.04 (-0.06 to 0.14) | 0.06 (-0.06 to 0.19) | 0.07 (-0.06 to 0.19) |
| **Height at age 7** | 0.12 (0.03 to 0.22) | 0.11 (0.02 to 0.21) | 0.13 (0.01 to 0.24) | 0.11 (0.00 to 0.22) |
| **Height at age 11** | 0.11 (0.01 to 0.20) | 0.08 (-0.02 to 0.18) | 0.10 (-0.01 to 0.21) | 0.09 (-0.03 to 0.20) |
| **Height at age 15** | 0.13 (0.03 to 0.22) | 0.10 (0.00 to -0.20) | 0.11 (0.00 to 0.22) | 0.10 (-0.02 to 0.21) |
| **Height at age 36** | 0.19 (0.10 to 0.28) | 0.18 (0.09 to 0.27) | 0.18 (0.07 to 0.28) | 0.18 (0.07 to 0.28) |
| **Height gain between 2 and 7** | 0.06 (-0.05 to 0.16) | 0.04 (-0.07 to 0.15) | 0.10 (-0.03 to 0.22) | 0.09 (-0.04 to 0.21) |
| **Height gain between 7 and 11** | 0.03 (-0.06 to 0.12) | 0.01 (-0.08 to 0.11) | 0.02 (-0.08 to 0.13) | 0.01 (-0.09 to 0.12) |
| **Height gain between 11 and 15** | 0.08 (-0.02 to 0.18) | 0.06 (-0.04 to 0.16) | -0.01 (-0.12 to 0.09) | -0.01 (-0.12 to 0.1) |
| **SITAR height size** | 0.15 (0.06 to 0.23) | 0.13 (0.04 to 0.22) | 0.09 (-0.01 to 0.2) | 0.09 (-0.02 to 0.19) |

***Supplementary Table 2:*** *Odds Ratio (OR) and 95% CI or p-values, respectively, of logistical GAMs of general health status at age of 69 and height loss. Model 1: univariate: only relative height loss, Model 2: multivariate: mutually adjusted for SITAR size height for overweight, education, region of birth, SEP in childhood, Model 3: additionally, mutually adjusted for smoking, fruit consumption and physical activity. Grey shaded cells show significant results.*

| | General health status (healthy vs less/not healthy) | | | | | | | |
|---|---|---|---|---|---|---|---|---|
| | **Model full sample** | | **Model 1** | | **Model 2** | | **Model 3** | |
| | **Men (N=838 )** | **Women (N=903)** | **Men (N=509)** | **Women (N=566)** | **Men (N=509)** | **Women (N=566)** | **Men (N=509)** | **Women (N=566)** |
| | **OR (95%CI) or p-value** | **OR (95%CI) or p-value** | **OR (95%CI) or p-value** | **OR (95%CI) or p-value** | **OR (95%CI) or p-value** | **OR (95%CI) or p-value** | **OR (95%CI) or p-value** | **OR (95%CI) or p-value** |
| **Relative height loss *** | p<0.001 | p<0.001 | p<0.001 | p=0.012 | p=0.001 | p=0.017 | p<0.001 | p=0.006 |
| **SITAR size height [$]** | | | | | p=0.509 | p=0.738 | p=0.631 | p=0.697 |
| **Overweight** | | | | | | | | |
| no | | | | | Ref. | Ref. | Ref. | Ref. |
| yes | | | | | 1.98 (1.26-3.12) | 1.60 (1.03-2.49) | 1.97 (1.25-3.10) | 1.62 (1.04-2.52) |
| **Education as adults** | | | | | | | | |
| no higher education | | | | | Ref. | Ref. | Ref. | Ref. |
| higher education | | | | | 0.95 (0.60-1.52) | 0.97 (0.66-1.42) | 1.00 (0.62-1.61) | 0.98 (0.66-1.45) |
| **SEP in childhood** | | | | | | | | |
| low SEP | | | | | Ref. | Ref. | Ref. | Ref. |
| medium SEP | | | | | 0.98 (0.59-1.65) | 0.99 (0.65-1.52) | 1.04 (0.61-1.75) | 1.02 (0.66-1.57) |
| high SEP | | | | | 1.12 (0.58-2.15) | 1.08 (0.63-1.84) | 1.17 (0.60-2.25) | 1.09 (0.64-1.86) |
| **Region of birth** | | | | | | | | |
| South | | | | | Ref. | Ref. | Ref. | Ref. |
| Middle | | | | | 0.83 (0.46-1.53) | 0.90 (0.57-1.41) | 0.81 (0.44-1.49) | 0.90 (0.57-1.42) |
| North | | | | | 1.04 (0.62-1.73) | 1.00 (0.65-1.52) | 1.01 (0.60-1.69) | 0.98 (0.64-1.50) |
| **Ever smoked** | | | | | | | | |
| no | | | | | | | Ref. | Ref. |
| yes | | | | | | | 1.55 (0.98-2.45) | 1.08 (0.74-1.58) |
| **Fruit consumption** | | | | | | | | |
| not every day | | | | | | | Ref. | Ref. |
| ever day | | | | | | | 1.05 (0.64-1.73) | 0.87 (0.56-1.36) |
| **Physical activity** | | | | | | | | |
| <1 / week | | | | | | | Ref. | Ref. |
| >=1 / week | | | | | | | 0.74 (0.47-1.16) | 1.27 (0.87-1.85) |

* smoothing parameter, probablities of height loss illustrated in Figure 2 (males) and Figure 3 (females)

[$] smoothing parameter, only p-values

***Supplementary Table 3:*** *Odds Ratio (OR) and 95% CI or p-values, respectively, of logistical GAMs of chronic disease score at age of 69 and height loss. Model 1: univariate: only relative height loss, Model 2: multivariate: mutually adjusted for SITAR size height for overweight, education, region of birth, SEP in childhood, Model 3: additionally, mutually adjusted for smoking, fruit consumption and physical activity. Grey shaded cells show significant results.*

| | **Chronic diseases** | | | | | | | |
|---|---|---|---|---|---|---|---|---|
| | **Model full sample** | | **Model 1** | | **Model 2** | | **Model 3** | |
| | **Men (N=937)** | **Women (N=967)** | **Men (N=532)** | **Women (N=584)** | **Men (N=532)** | **Women (N=584)** | **Men (N=532)** | **Women (N=584)** |
| | **OR (95%CI) or p-value** | **OR (95%CI) or p-value** | **OR (95%CI) or p-value** | **OR (95%CI) or p-value** | **OR (95%CI) or p-value** | **OR (95%CI) or p-value** | **OR (95%CI) or p-value** | **OR (95%CI) or p-value** |
| **Relative height loss *** | p=0.006 | p=0.003 | p=0.023 | p=0.208 | p=0.047 | p=0.219 | p=0.055 | p=0.136 |
| **SITAR size height** $^{\$}$ | | | | | p=0.720 | p=0.893 | p=0.817 | p=0.877 |
| **Overweight** | | | | | | | | |
| no | | | | | Ref. | Ref. | Ref. | Ref. |
| yes | | | | | 2.82 (1.72-4.61) | 1.56 (0.94-2.59) | 2.89 (1.75-4.76) | 1.59 (0.95-2.64) |
| **Education as adults** | | | | | | | | |
| no higher education | | | | | Ref. | Ref. | Ref. | Ref. |
| higher education | | | | | 0.93 (0.57-1.54) | 1.17 (0.74-1.85) | 0.97 (0.58-1.61) | 1.21 (0.76-1.92) |
| **SEP in childhood** | | | | | | | | |
| low SEP | | | | | Ref. | Ref. | Ref. | Ref. |
| medium SEP | | | | | 1.36 (0.77-2.41) | 1.09 (0.65-1.81) | 1.51 (0.84-2.70) | 1.15 (0.68-1.92) |
| high SEP | | | | | 1.26 (0.61-2.59) | 0.92 (0.48-1.76) | 1.38 (0.66-2.86) | 0.92 (0.47-1.77) |
| **Region of birth** | | | | | | | | |
| South | | | | | Ref. | Ref. | Ref. | Ref. |
| Middle | | | | | 0.90 (0.48-1.70) | 0.87 (0.51-1.51) | 0.86 (0.46-1.63) | 0.88 (0.51-1.52) |
| North | | | | | 0.77 (0.44-1.35) | 1.12 (0.68-1.85) | 0.73 (0.42-1.29) | 1.09 (0.66-1.8) |
| **Ever smoked** | | | | | | | | |
| no | | | | | | | Ref. | Ref. |
| yes | | | | | | | 1.59 (0.96-2.61) | 1.14 (0.73-1.78) |
| **Fruit consumption** | | | | | | | | |
| not every day | | | | | | | Ref. | Ref. |
| ever day | | | | | | | 1.13 (0.66-1.94) | 0.66 (0.4-1.1) |
| **Physical activity** | | | | | | | | |
| <1 / week | | | | | | | Ref. | Ref. |
| >=1 / week | | | | | | | 0.57 (0.34-0.93) | 1.51 (0.96-2.36) |

* smoothing parameter, probablities of height loss illustrated in Figure 2 (males) and Figure 3 (females)

$^{\$}$ smoothing parameter, only p-values

***Supplementary Table 4:*** *Odds Ratio (OR) and 95% CI or p-values, respectively, of logistical GAMs of osteoarthritis between 60 and 69 years and height loss. Model 1: univariate: only relative height loss, Model 2: multivariate: mutually adjusted for SITAR size height for overweight, education, region of birth, SEP in childhood, Model 3: additionally, mutually adjusted for smoking, fruit consumption and physical activity. Grey shaded cells show significant results.*

| | **Osteoarthritis** | | | | | | | |
|---|---|---|---|---|---|---|---|---|
| | **Model full sample** | | **Model 1** | | **Model 2** | | **Model 3** | |
| | **Men (N=930)** | **Women (N=967)** | **Men (N=534)** | **Women (N=584)** | **Men (N=534)** | **Women (N=584)** | **Men (N=534)** | **Women (N=584)** |
| | **OR (95%CI) or p-value** | **OR (95%CI) or p-value** | **OR (95%CI) or p-value** | **OR (95%CI) or p-value** | **OR (95%CI) or p-value** | **OR (95%CI) or p-value** | **OR (95%CI) or p-value** | **OR (95%CI) or p-value** |
| **Relative height loss *** | p=0.001 | p=0.011 | p=0.001 | p=0.094 | p=0.004 | p=0.151 | p=0.002 | p=0.099 |
| **SITAR size height [$]** | | | | | p=0.384 | p=0.200 | p=0.362 | p=0.212 |
| **Overweight** | | | | | | | | |
| no | | | | | Ref. | Ref. | Ref. | Ref. |
| yes | | | | | 2.57 (1.51-4.38) | 1.61 (1.02-2.53) | 2.59 (1.51-4.45) | 1.61 (1.02-2.55) |
| **Education as adults** | | | | | | | | |
| no higher education | | | | | Ref. | Ref. | Ref. | Ref. |
| higher education | | | | | 0.94 (0.55-1.63) | 1.33 (0.89-2.00) | 0.98 (0.56-1.72) | 1.32 (0.88-1.99) |
| **SEP in childhood** | | | | | | | | |
| low SEP | | | | | Ref. | Ref. | Ref. | Ref. |
| medium SEP | | | | | 0.88 (0.47-1.64) | 1.16 (0.74-1.82) | 0.87 (0.46-1.64) | 1.16 (0.74-1.83) |
| high SEP | | | | | 1.47 (0.71-3.06) | 1.05 (0.59-1.86) | 1.49 (0.71-3.15) | 1.06 (0.59-1.88) |
| **Region of birth** | | | | | | | | |
| South | | | | | Ref. | Ref. | Ref. | Ref. |
| Middle | | | | | 0.41 (0.18-0.94) | 1.04 (0.65-1.66) | 0.41 (0.18-0.95) | 1.04 (0.65-1.66) |
| North | | | | | 0.98 (0.55-1.74) | 0.83 (0.53-1.31) | 1.01 (0.56-1.80) | 0.83 (0.52-1.31) |
| **Ever smoked** | | | | | | | | |
| no | | | | | | | Ref. | Ref. |
| yes | | | | | | | 1.97 (1.14-3.43) | 1.03 (0.69-1.53) |
| **Fruit consumption** | | | | | | | | |
| not every day | | | | | | | Ref. | Ref. |
| ever day | | | | | | | 2.06 (1.08-3.93) | 1.16 (0.71-1.90) |
| **Physical activity** | | | | | | | | |
| <1 / week | | | | | | | Ref. | Ref. |
| >=1 / week | | | | | | | 1.50 (0.88-2.58) | 0.96 (0.64-1.45) |

* smoothing parameter, probablities of height loss illustrated in Figure 2 (males) and Figure 3 (females)

[$] smoothing parameter, only p-values

***Supplementary Table 5:*** *Odds Ratio (OR) and 95% CI or p-values, respectively, of logistical GAMs of Pain while walking at age of 69 and height loss. Model 1: univariate: only relative height loss, Model 2: multivariate: mutually adjusted for SITAR size height for overweight, education, region of birth, SEP in childhood, Model 3: additionally, mutually adjusted for smoking, fruit consumption and physical activity. Grey shaded cells show significant results.*

| | Pain while walking | | | | | | | |
|---|---|---|---|---|---|---|---|---|
| | Model full sample | | Model 1 | | Model 2 | | Model 3 | |
| | Men (N=803) | Women (N=863) | Men (N=531) | Women (N=577) | Men (N=531) | Women (N=577) | Men (N=531) | Women (N=577) |
| | OR (95%CI) or p-value | OR (95%CI) or p-value | OR (95%CI) or p-value | OR (95%CI) or p-value | OR (95%CI) or p-value | OR (95%CI) or p-value | OR (95%CI) or p-value | OR (95%CI) or p-value |
| **Relative height loss *** | p=0.001 | p<0.001 | p=0.012 | p=0.004 | p=0.048 | p=0.043 | p=0.052 | p=0.041 |
| **SITAR size height $^{\$}$** | | | | | p=0.188 | p=0.087 | p=0.172 | p=0.085 |
| **Overweight** | | | | | | | | |
| no | | | | | Ref. | Ref. | Ref. | Ref. |
| yes | | | | | 1.47 (0.92-2.35) | 2.72 (1.71-4.34) | 1.47 (0.92-2.34) | 2.86 (1.78-4.58) |
| **Education as adults** | | | | | | | | |
| no higher education | | | | | Ref. | Ref. | Ref. | Ref. |
| higher education | | | | | 0.55 (0.34-0.89) | 1.11 (0.72-1.73) | 0.57 (0.35-0.93) | 1.16 (0.74-1.82) |
| **SEP in childhood** | | | | | | | | |
| low SEP | | | | | Ref. | Ref. | Ref. | Ref. |
| medium SEP | | | | | 1.01 (0.59-1.73) | 0.76 (0.47-1.23) | 1.04 (0.60-1.78) | 0.81 (0.49-1.32) |
| high SEP | | | | | 1.03 (0.52-2.05) | 0.74 (0.40-1.37) | 1.07 (0.54-2.13) | 0.82 (0.44-1.52) |
| **Region of birth** | | | | | | | | |
| South | | | | | Ref. | Ref. | Ref. | Ref. |
| Middle | | | | | 0.45 (0.22-0.90) | 0.85 (0.52-1.39) | 0.44 (0.22-0.89) | 0.90 (0.55-1.48) |
| North | | | | | 1.02 (0.61-1.69) | 0.57 (0.34-0.95) | 1.00 (0.60-1.67) | 0.54 (0.32-0.90) |
| **Ever smoked** | | | | | | | | |
| no | | | | | | | Ref. | Ref. |
| yes | | | | | | | 1.31 (0.82-2.09) | 1.91 (1.25-2.93) |
| **Fruit consumption** | | | | | | | | |
| not every day | | | | | | | Ref. | Ref. |
| ever day | | | | | | | 0.87 (0.53-1.42) | 1.34 (0.79-2.29) |
| **Physical activity** | | | | | | | | |
| <1 / week | | | | | | | Ref. | Ref. |
| >=1 / week | | | | | | | 0.88 (0.55-1.40) | 1.2 (0.77-1.86) |

* smoothing parameter, probablities of height loss illustrated in Figure 2 (males) and Figure 3 (females)

$^{\$}$ smoothing parameter, only p-values
